# A cross-sectional survey of the workplace factors contributing to symptoms of anxiety and depression among nurses and physicians during the first wave of COVID-19 pandemic in two US healthcare systems

**DOI:** 10.1101/2021.01.25.21250315

**Authors:** Igor Burstyn, Karyn Holt

## Abstract

**Background:** Anxiety and depression among physicians and nurses during COVID-19 pandemic in the USA is not well described and its modifiable causes poorly understood.

**Methods:** We conducted a cross-sectional survey of symptoms of anxiety and depression (Hospital Anxiety and Depression Scale) among physicians and nurses in two US healthcare systems June-Sept 2020. We ascertained features of work as well as its perceptions and associated concerns in relation to risk of anxiety and depression, while controlling for health history via regression and path analyses.

**Results:** About a third of 684 nurses and 185 physicians surveyed showed symptoms of anxiety or depression, the excess was particularly prominent in nurses. Belief in having been infected was a dominant cause of anxiety and depression, more related to history of symptoms of pneumonia, then the contact with infected patients. Having confidence in competent use and access to personal protective equipment, maintaining usual working hours and being surrounded by colleagues who were both sufficient in numbers and not stressed, was protective. Having support of immediate family and religious communities lessened anxiety and depression after accounting for other factors. Involvement in aerosol-generating procedures with infected patients was linked with lower depression in nurses but higher among physicians. Likewise, the setting of recent patient encounters affected risk differently for physicians and nurses.

**Conclusions:** Our findings may help develop mitigation measures and underscore the need to help nurses and physicians bear the psychological burden of COVID-19 pandemic and similar events in the future.

## Introduction

Healthcare workers (HCW) are presumed to be at risk for nosocomial infections with SARS-CoV-2, virus causing COVID-19 disease. There is robust and widely publicized evidence that HCWs in the US are at increased risk. According to the US CDC, 55% of HCW with COVID-19 reported contact with infected individuals only in a healthcare setting and they were the dominant occupational group among diagnosed cases during the onset of the epidemic.^1^ The infection rate for SARS-CoV-2 in the early days of the epidemic was 7.3% in one study of the US HCWs while only affecting 0.4% of others,^2^ and continued to increase, as the epidemic progressed towards its second wave, among HCWs, with nurses being reported as having the highest infection rates.^3^

Given that HCWs understand better than most that they are at an elevated risk of any infection during an outbreak of a novel infection, they can be expected to be at risk of psychological distress, whether they themselves become infected as was the cases during SARS epidemic.^4, 5^ For example, Chong et al.,^6^ reported pervasive emotional distress, feelings of extreme vulnerability, uncertainty, and threats to life, during the rapid spread of SARS. Soklaridis et al.^7^ argued that review of evidence available as of June 2020 indicates that HCWs in general are among those who are particularly distressed and fearful during pandemics, aggravated by many factors, including concerns about workload, exposure, shortages of personal protective equipment (PPE), and inadequate support. The authors identified lack of consideration of pre-existing medical conditions as one of the weaknesses in available evidence and emphasized that cultural context must be considered, implying the need for local data to inform mediation measures. Synthesis of relevant literature on COVID-19 and earlier similar outbreaks by Preti et al.^8^ reveals elevated anxiety and depression among HCWs, mitigate by plethora of work-related factors (including support and confidence in PPE), some presumably modifiable. With the SARS outbreak, new onset mental ill-health was no greater, one year later, in HCW involved in the care of SARS patients than rates in the community,^9^ but that may not be a valid prediction of effects with the COVID-19 pandemic which has had longer disruptive effects on the lives of a greater number of HCWs, both in breadth and depth.

Meta-analysis of studies from China and Singapore by Pappa et al. ^10^ suggests high levels of anxiety and depression among HCW involved in the care of patients with COVID-19 early in the epidemic, with somewhat higher levels among nurses (26-30%) compared to physicians (22-25%); the risk was higher on average among female HCWs. In one study included in the meta-analysis, Lai et al.^11^ reported that among 1,257 HCWs in 34 hospitals in China, during January-February 2020, the symptoms of anxiety and depression (exhibited by more about half of the participants) were elevated on average by 50% among those who were engaged in direct care of COVID-19 patients; higher known infection rates in a region where HCWs practiced adversely affected mental health. Likewise, in the largest study included in the meta-analysis of 11,118 HCWs in China,^12^ authors reported that among 3,351 frontline HCWs there was on average doubling of “severe” anxiety and depression compare to non-frontline HCWs. Wang et al.^13^ observed that poor self-rated health, having a chronic illness, suspected contact with infected person (but not confirmed case) and specific symptoms of ill-health consistent with COVID-19 during previous 14 days were associated with elevated symptoms of anxiety and depression in a general population sample in China of 1,210 respondents from 194 cities collected in February 2020, suggesting that the same associations may also exist among HCWs. Perception of lack of adequacy of PPE and infection control during COVID-19 epidemic was associated with increased symptoms of anxiety and depression among 5,988 Canadian HCWs during spring of 2020.^14^

In the US,^15^ a nation-wide convenience sample (high in emergency department staff) of 2,040 HCWs during May 2020 revealed that having reported symptoms consistent with COVID-19 was associated with anxiety and depression. It must be noted that almost a third of the participants were suspected of having COVID-19 (a far higher rate than expected from a random sample at the time), further limiting works’ generalizability; pre-existing anxiety, depression and perception of mental health support were not evaluated. First responders, including 98 hospital staff, from Rocky Mountain region of the US during spring of 2020,^16^ exhibited evidence of excess of anxiety and depression due to contact with COVID-19 patients and their own reported immunocompromised status. Czeisler et al.^17^ provide evidence of increase in anxiety and depression in the US in general during April-June 2020 compared to the same period a year before, with a notable excess of having considered suicide among essential workers (who presumably include HCWs); the survey highlighted importance of adjusting for history of anxiety and depression, including whether it was recently treated. Overall, data on anxiety and depression among HCWs in US during COVID-19 epidemic appears to be limited, with few indications of whether modifiable causes seen in other populations are at play.

We aimed to identify workplace factors that place physicians and nurses at risk for anxiety and depression during the first wave of the COVID-19 pandemic in samples from two healthcare systems, accounting for health history, perceived risks, and support.

## Materials and Methods

We designed a cross-sectional survey of all physicians and nurses employed and contracted by the Tower Health in Southeastern Pennsylvania (TH) and the University Medical Center, Las Vegas, Nevada (UMC), and licensed to practice in these states, during the spring and summer of 2020, corresponding to the early phases of the COIVD-19 epidemic in the US. Participation was both voluntary and anonymous, unless the participants chose to enter their name in the survey wishing to be contacted for follow-up (yet to be conducted). TH is regional healthcare provider that offers healthcare and wellness services to a population of 2.5 million people in Philadelphia and Southeastern Pennsylvania. It includes six acute care hospitals and other entities that provide a full range of medical care, wellness programs, and public health services. TH consists of numerous hospitals, including a pediatric hospital, a partnership with Drexel University, in Philadelphia, home healthcare services, and a network of 22 urgent care facilities. The UMC is a government hospital and the only level one trauma center in Las Vegas, with 564 total hospital beds. It is the eighteenth largest public hospital in the US, providing both adult and pediatric care over portions of Nevada, California, Arizona, and Utah.

We collected data via an online survey (implemented in Qualtrics hosted by Drexel University). The invitation to enroll in the study was distributed by emails, using mailing lists held by TH and UMC, containing links to online surveys. The initial recruitment email was sent out followed by reminder emails, one week apart for a total of a four-week periods. Ethics approvals were obtained from Drexel University and the University of Nevada, Las Vegas for TH and UMC sites, respectively.

We were primarily interested in information on work conditions and personal health since the start of the pandemic, defined by dates when the first cases of COVID-19 were reported in each state: March 10 for TH and March 5 for UMC. Some questions concerned the most recent week worked since diagnosis of the first case in each state. On June 3, 2020, we distributed invitation to TH survey aimed at nurses to advanced nursing practitioners (203) and registered nurses (4,336); at the same time, we distributed invitation to TH survey aimed at physicians to 2,496 active medical staff and 204 physician assistants; all messages were delivered to the recipients. On September 9, 2020, we distributed invitations to the UMC version of the survey to both nurses (1518) and physicians (1186).

We used the *Hospital Anxiety and Depression Scale* (HADS), to measure symptoms of anxiety and depression separately; scores of equal to or above 11 indicate presence of these conditions but are not equivalent to clinical diagnosis.^18, 19^ Higher scores indicated higher chance of having the conditions. The *Community-Acquired Pneumonia Symptom Questionnaire* (CAP-Sym) uses standard list of symptoms of wide range of infections to determine risk of pneumonia, symptomatically close to COVID-19. Lamping et al.^20^ developed and validated the instrument. We used CAP-Sym to determine whether our participants experience symptoms consistent with COVID-19 since start of the epidemic in each state. We evaluated resilience using the two-item Connor Davidson Resilience Scale (CD-RISC2).^21^

In addition to demographic characteristics (age, marital status, years in profession, gender, educational level, and location of unit and duty assignment), we queried contact with known or suspected COVID-19 patients, involvement with aerosol-generating procedures on known and suspected COVID-19 patients (suspected risk of infection at the time and thus a plausible source of anxiety), belief about having been infected with virus that causes COVID-19, history of anxiety and depression prior to the epidemic (and evidence of is exacerbation requiring treatment in a year before the epidemic), history of respiratory and other conditions known at the time to place person at elevated risk of death due to COVID-19 (asthma, chronic obstructive pulmonary disease, emphysema: modeled on Canadian Community Health Survey elements,^22^ as well as a battery of questions about perceptions (captured on Likert-like scale ranging from 0 to 100) of working conditions in most recent week of work, confidence in work safety (including personal protective equipment (PPE), source of anticipated support during pandemic, and specific worries. “Worrying” is an established proximal antecedent of generalized anxiety (such as assessed by HADS) as opposed to a more distal “environmental” cause.^23, 24^ Consequently, we did not adjust for worries in regression models of HADS scores described below, but rather (a) investigated association between worries and HADS for anxiety in principal components analysis and (b) used reported worries descriptively with respect to their correlation with HADS scores. Copies of research instruments are available upon request, but the key questions not present in the cited literature are reported as part of results below.

All calculations were performed in SAS v 9.4 (SAS Institute, Cary, NC). Association of HADS scores for anxiety (HADS A) and depression (HADS D) were examined for each of the covariate of interest in terms of counts of scores ≥11 (referred to as “cases” hereafter) for categorical covariates and Spearman rank correlations for continuous covariates. Univariate associations of continuous HADS scores with categorical variables was evaluated in Kruskal-Wallis (K-W) tests. We conducted path analysis to determine relationships between HADS scores, belief in having been infected, history of symptoms on pneumonia since start of infection (CAP-Sym) and belief about contact with COVID-19 patients (PROC CALIS … method=MLM).^25^ All analyses were stratified by profession (nurse vs. physician) and study site (TH vs. UMC); we chose not to pool data to preserve unique features of each site and profession. Multivariable regression models of HADS scores were estimated using binomial regression on TH data only (it proved to be of sufficient size to yield stable regression models that converged; PROC GENMOD). These yielded relative rates (RR) and 95% confidence intervals (CI) of change in HADS scores in relation to variables that showed evidence of association with HADS scores in univariate analyses, adjusted for each other and plus all demographic variables. Missing values of continuous variables were replaced with means of observed values.

## Results

### Nurses: demographics, work, and health histories

Nurses recruited at TH (623) and UMC (61) shared many characteristics in terms of demographics and levels of symptoms of anxiety, depression, and history of episodes of pneumonia since onset of the epidemic. Few of them were tested for COVID-19: among 43 tested TH nurses, 10 were positive, and among 12 tested UMC nurses 6 were positive (**Table I**). Prevalence of anxiety cases (34% at TH and 39% at UMC) exceeded that of depression cases (12% at TH and 11% at UMC). Depression and anxiety scores had rank correlation of 0.7 (p<0.0001) in both groups of nurses.

**Table I:**
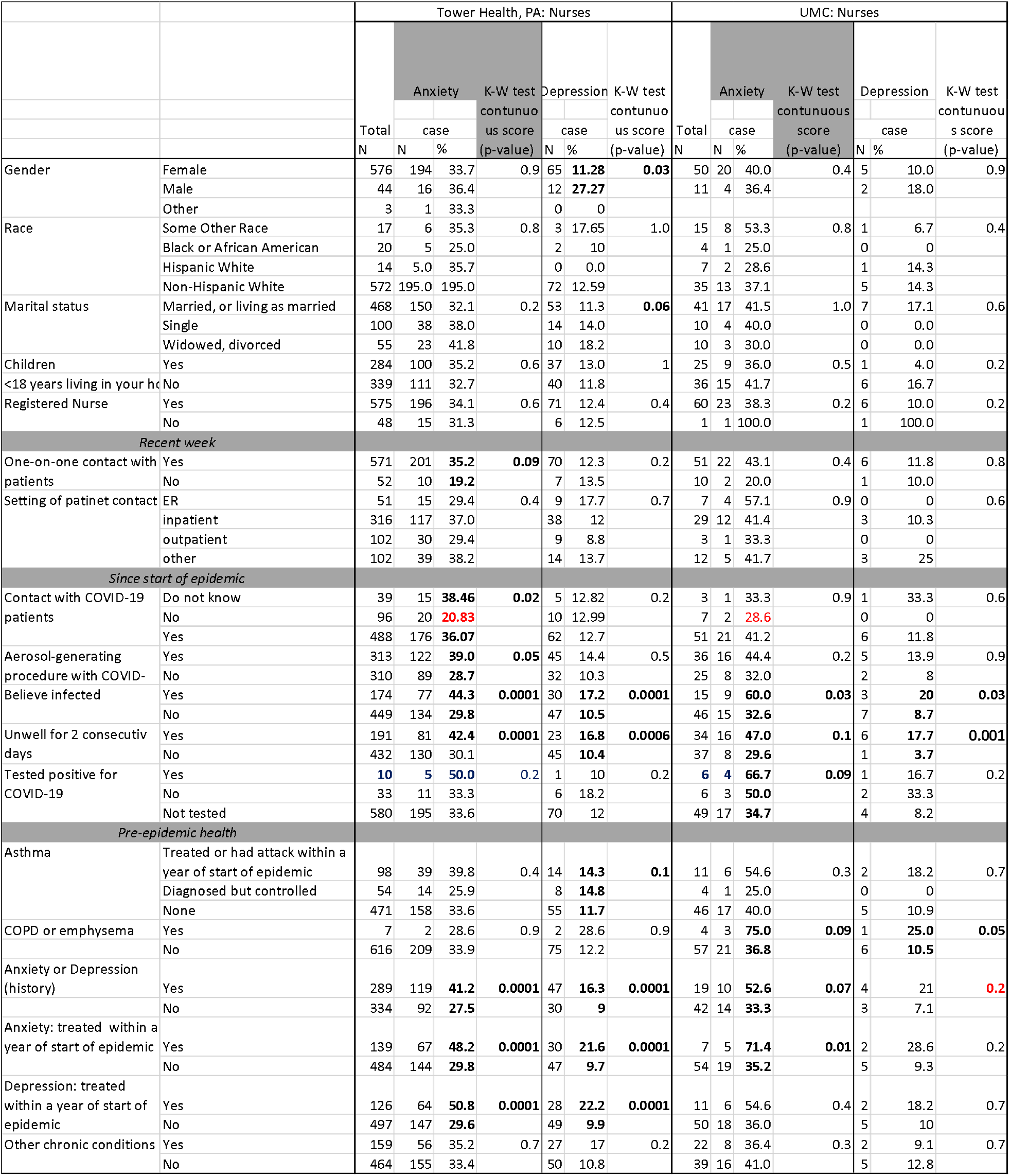
Description of *nurses* and differences in their HADS scores: cases with Hospital Anxiety Depression Scale (HADS) scores ≥11 and continuous scores (Kruskal-Wallis tests, K-W)

The enrolled TH nurses were aged 21 to 70 with average of 43 (SD 12) years; they qualified between 1975 and 2020, with the mean year of qualification of 2003 (SD 12). They had an average HADS anxiety score of 8.7 (SD 4.6), with 5^th^, 50^th^ and 95^th^ percentiles of 1, 8, and 17, respectively. TH nurses had an average HADS depression score of 5.7 (SD 4.0), with 5^th^, 50^th^ and 95^th^ percentiles of 0, 5, and 12, respectively. Among TH nurses, CAP-Sym scores was on average 12 (SD 19.2) and was weakly correlated with both HADS scores (0.2, p<0.0001), and CD-RISC2 was on average 6 (SD 1) and inversely related to HADS scores (−0.4, p<0.0001).

The enrolled UMC nurses were aged 25 to 67 with average of 46 (SD 11) years; they qualified between 1979 and 2019, with the mean year of qualification of 2002 (SD 10). They had an average HADS anxiety score of 9.4 (SD 4.6), with 5^th^, 50^th^ and 95^th^ percentiles of 2, 9, and 17, respectively. UMC nurses had an average HADS depression score of 6.2 (SD 3.9), with 5^th^, 50^th^ and 95^th^ percentiles of 0, 7, and 12, respectively. Among UMC nurses, CAP-Sym scores was on average 22 (SD 21.2) and was positively correlated with HADS scores: 0.3 with anxiety scored (p=0.04) and 0.5 with depression score (p=0.0002). The CD-RISC2 was on average 6 (SD 1) and inversely related to HADS scores (−0.3, p<0.02).

Nurses were predominantly female and non-Hispanic White, who were married and had children under 18 years of age living at home; the majority were registered nurses (**Table I**). We noted evidence of excess of cases of anxiety among TH nurses who had recent direct patient contact within a week (35% vs. 19%) or were involved in aerosol-generating procedures during epidemic (39% vs 29%). We saw lower rates of anxiety among nurses who did not think that they had contact with COVID-19 patients (21% vs 36-38%). The evidence for these tendencies is supported by low p-values of K-W tests in the TH sample. Nurses we surveyed at UMC exhibited similar patterns. There was little evidence of such association with HADS depression scores, either continuous or dichotomized at ≥11. There was no evidence of association with setting of patient contact in the most recent week of work.

Among health-related factors, one of the most striking features of the data, consistent across study sites, is the higher rates of anxiety and depression cases among nurses who were unwell for two consecutive days since start of the epidemic and who believed that they were infected (**Table I**). For example, among TH nurses those who believed they were infected (n=174) had rate of anxiety (44%) and depression (17%), compared to corresponding rates of 30% and 11% among 449 nurses who did not believe to have been infected. History of anxiety and depression, especially those requiring treatment were also associated with elevated prevalence cases of both anxiety and depression, but no such pattern was seen for respiratory disease. The patterns of results were similar for both groups of nurses.

### Physicians: demographics, work, and health histories

We recruited 135 physicians at TH and 50 at UMC (50). Physicians in TH and UMC samples were mostly non-Hispanic white and married, with about half reporting that they had children under 18 living at home (**Table II**); sample of TH physicians was gender-balanced, but there were more men in the UMC sample. Just as with nurses, the majority were not tested for COVID-19: 1 of 9 positive among TH physicians, and 3 out of 8 positives among UMC physicians. As with nurses, among physicians the prevalence of anxiety cases (19% at TH and 12% at UMC) exceeded that of depression cases (5% at TH and 4% at UMC). Depression and anxiety scores had rank correlation of 0.7-0.8 (p<0.0001) among physicians. The rates of anxiety and depression appear to be lower among physicians than among nurses at both sites.

**Table II:**
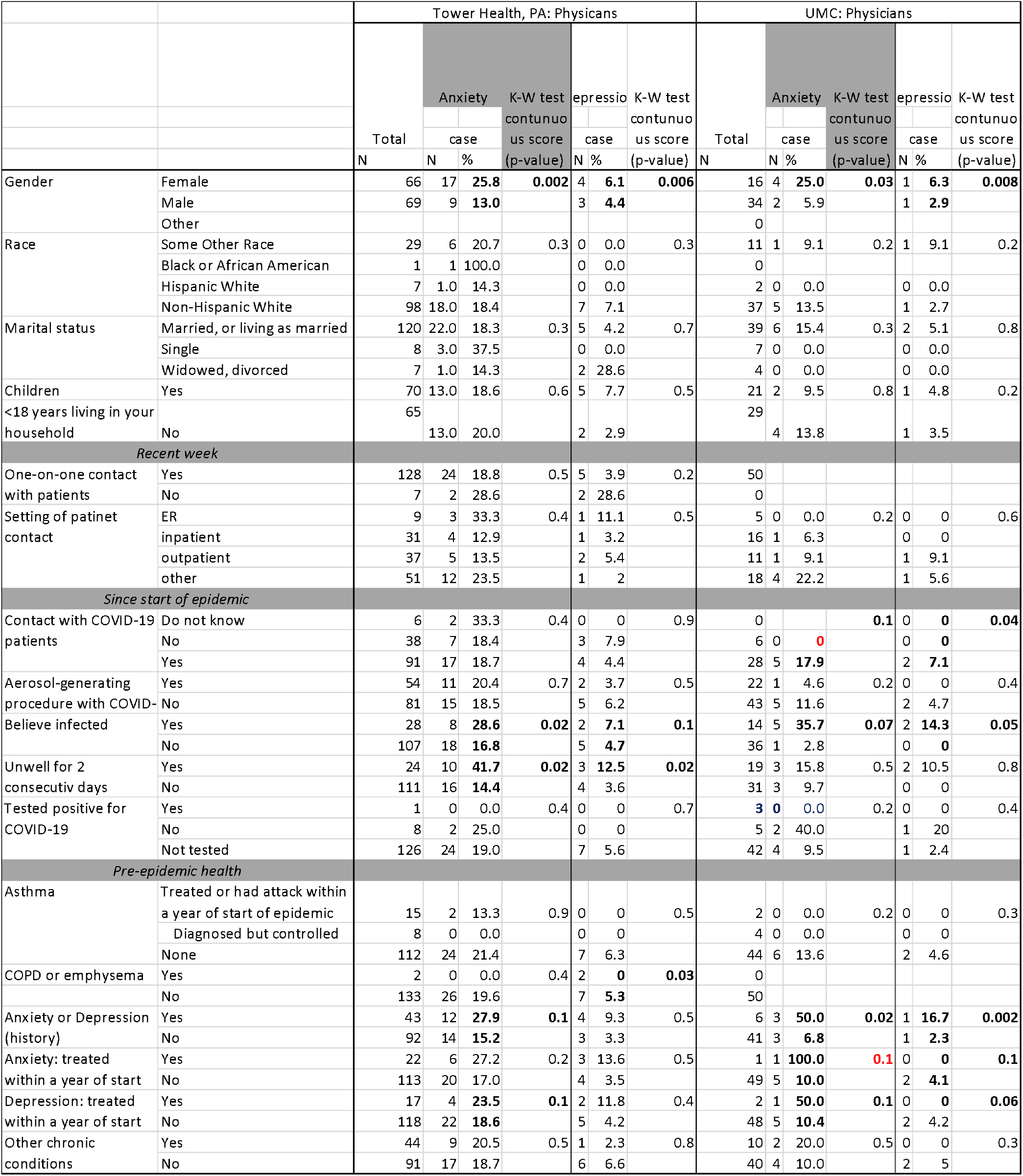
Description of *physicians* and differences in their HADS scores: cases with Hospital Anxiety Depression Scale (HADS) scores ≥11 and continuous scores (Kruskal-Wallis tests, K-W)

Physicians were somewhat older than nurses: TH physicians were on average 49 (SD 12) years of age, UMC physicians – 52 (SD 11). The physicians qualified between 1970 and 2019, medians in the late 1990’s. Physicians had lower HADS and CAP-Sym scores than nurses in their respective healthcare systems, as detailed below.

TH physicians had an average HADS anxiety score of 7.1 (SD 4.0), with 5^th^, 50^th^ and 95^th^ percentiles of 1, 7, and 14, respectively. They had an average HADS depression score of 4.2 (SD 3.4), with 5^th^, 50^th^ and 95^th^ percentiles of 0, 4, and 11, respectively. Among TH physicians, CAP-Sym scores was on average 5.6 (SD 12.8) and correlated with both HADS scores (r=0.2, p=0.01). The CD-RISC2 was on average 7 (SD 1) and inversely related to HADS scores (r=-0.3, p<0.001).

UMC physicians had lower HADS scores than their TH colleagues. Specifically, their average HADS anxiety score of 5.4 (SD 3.9), with 5^th^, 50^th^ and 95^th^ percentiles of 0, 5, and 13, respectively. They had an average HADS depression score of 3.9 (SD 4.0), with 5^th^, 50^th^ and 95^th^ percentiles of 0, 2.5, and 10, respectively. Among UMC physicians, CAP-Sym scores was on average 12.4 (SD 17.4), higher than at TH; it was not correlated with HADS scores (r=0.1, p>0.4). The CD-RISC2 was on average 7 (SD 1) and inversely related to HADS scores for anxiety (r=-0.6) and depression (r=-0.5) (p<0.0001).

The belief that physicians were infected was associated with elevated rate of anxiety and depression cases among all physicians. Having been unwell for two consecutive days since start of the epidemic likewise was associated with higher HADS scores among TH but not UMC physicians (although sample is small). Unlike with nurses, there appears to be no evidence of excess of cases of anxiety among TH physicians who had recent direct patient contact during the most recent week of work; all UMC physicians had such patient contact. There was no evidence of association of HADS scores with having been involved in aerosol-generating procedures during epidemic or setting of recent contact with the patients. There is a suggestion of elevated HADS scores among UMC but not TH physicians who reported that they had contact with COVID-19 patients, but it is based on small numbers. History of anxiety and depression, but not other conditions, as with nurses, were related to higher prevalence of cases of anxiety and depression.

### Concerns and perceptions

Univariate analysis of concerns and perception in relation to HADS scores is presented in **Table III** for nurses and in **Table IV** for physicians.

**Table III:**
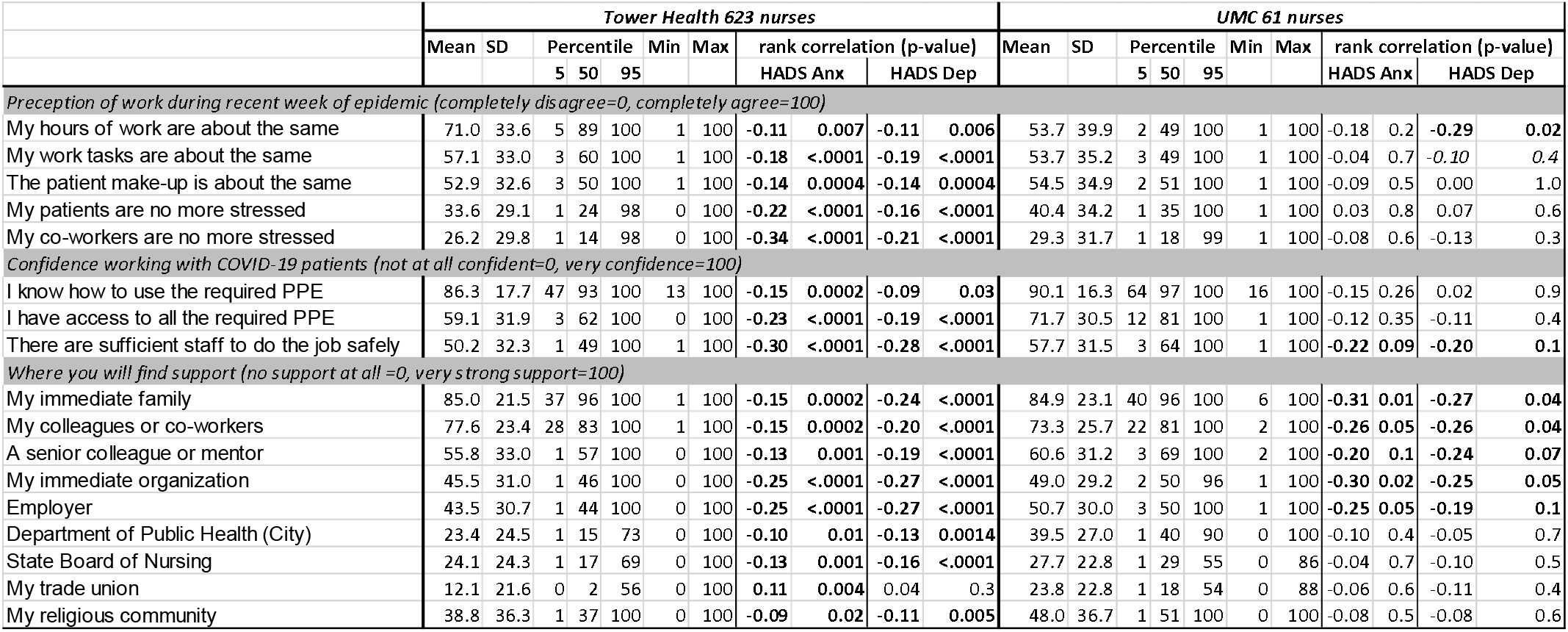
Perceptions and concerns during COVID-19 epidemic of *nurses* in relation to Hospital Anxiety Depression Scale (HADS) scores for anxiety (Anx) and depression (Dep).

**Table IV:**
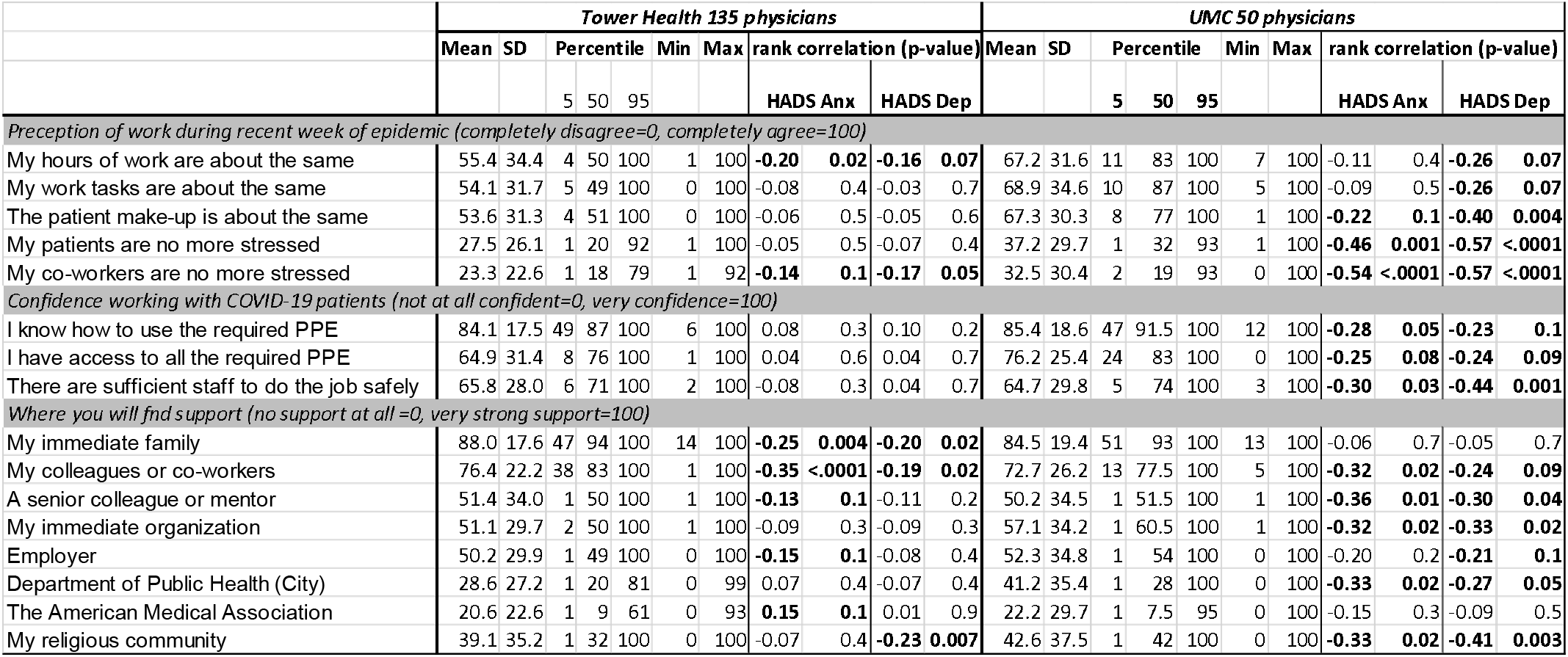
Perceptions and concerns during COVID-19 epidemic of *physicians* in relation to Hospital Anxiety Depression Scale (HADS) scores for anxiety (Anx) and depression (Dep).

When asked to record “perception of work during recent week of epidemic” on a Likert-like scale ranging from (completely disagree=0) to (completely agree=100), nurses tended to agree that “hours of work”, “tasks” and “patient make-up” did not change, with median scores of at or above 50 (**Table III**). On the other hand, both patients and co-workers were perceived as more stressed, with most tending to disagree that this was so, with median scores <35. All these perceptions were negatively correlated with HADS scores among TH nurses, with the lower levels of anxiety and depression associated with lower perceived stress among co-workers during recent week of work, rank correlations of −0.34 and - 0.21 (p<0.0001) for anxiety and stress, respectively. Among UMC nurses, only the perception that working hours were about the same during recent week of work was associated with reduced HADS scores, i.e., lower risk, with rank correlations of −0.18 (p=0.2) and −0.29 (p=0.02) for anxiety and depression, respectively. Among physicians, we observed similar patterns to those among nurses (**Table IV**). However, we noted inverse relationships with HADS scores among UMC physicians with “the patient make-up is about the same”, “my patients are no more stressed” and “my co-workers are no more stressed.” Association of lower HADS scores with “working ours being the same” and less stress among co-workers (but not other perceptions) was evident among TH physicians.

Confidence in working with COVID-19 patients with respect to PPE use and sufficient staffing was high among nurses, with median scores above 50 and confidence in knowledge of how to use of PPE reaching median scores of >90 on a scale of (not at all confident=0) to (very confidence=100) (**Table III**).

Confidence in “sufficient staff to do the job safely” was most strongly protective against anxiety and depression, with rank correlations of −0.3 (p<0.0001) in TH and −0.2 (p=0.1) in UMC cohorts. Among physicians, confidence in PPE and staffing was likewise high, but its protective effects appeared to be limited to UMC physicians (**Table IV**).

Nurses at TH tended to report finding strong support only among immediate family, colleagues or co-workers, or a senior colleague or mentor, with median scores at or above 50 on a scale of (no support at all =0) to (very strong support=100) (**Table III**). Among TH nurses, greater confidence in *any* source of support was associated with reduction of HADS scores (lower risk), except for the reverse trend with reports of finding support from the trade union at TH: r=0.1, p=0.004 for anxiety and r=0.04, p=0.3 for depression. Nurses at UMC tended to believe that they would find stronger support among immediate family, colleagues or co-workers, a senior colleague or mentor, their immediate organization, and employer, with median scores at or above 50 on a scale of (no support at all =0) to (very strong support=100). Only support from these sources was associated with lower HADS scores (reduced risk) in among UMC nurses, with rank correlations of −0.2 to −0.3 (p≤0.1). On average, nurses did not expect to find support from municipal department of public health, State Boards of Nursing, and trade unions, with median scores <20 in TH cohort and 20-40 in UMC cohort.

Among physicians, the dominant reported sources of support were the same as among nurses, with reports of the perceived strongest support from immediate family (average scores >80) and perception of American Medical Association being the least likely source of support (average scores around 20) (**Table IV**). Perception of stronger support from colleagues and co-workers was associated with lower HADS scores in both groups of physicians, e.g., for anxiety r=-0.35, p<0.0001 at TH and −0.32, p=0.02 at UMC. Lower depression scores were related to perception of stronger support from religious communities among physicians (r=-0.23, p=0.007 at TH, −0.41, p=0.003 at UMC). There was a tendency for perception of stronger support from any source other than immediate family, to be linked to lower HADS scores among UMC physicians. The pattern was different for TH physicians for whom perception of stronger support from family appeared to be protective (r=-0.25, p=0.004 and r=-0.02, p=0.02 for anxiety and depression, respective), while most perceptions appeared to not be strongly associated with HADS scores.

We inquired about “worries about the COVID-19 epidemic” and captured it on a Likert-like scale ranging from (not at all worried=0) to (very worried=100). As illustrated in **Table V**, by far the greatest worry with an average scores 70 and above was of infecting one’s family, followed by worry about being infected oneself (average scores above or near 60). Worries related to performance of professional duties were relatively less prominent, e.g., with respect to adequacy of experience and not being able to cope the scores were in the 30’s and 40’s on average, with nurses being somewhat more concerned than physicians. These patterns were consistent across sites and professional groups. The strongest of the associations with HADS scores, for each site and profession, was a worry that the person will fail themselves and their family. HADS scores for anxiety and all responses about worries were associated with one latent component in principal components analysis accounting for the majority of common variance (e.g., 50% among TH nurses, the most informative of our samples, and 43% for TH physicians); only one latent component was suggested by the scree plots (details not shown).

**Table V:** Reported worries about the COVID-19 epidemic among Tower Health (TH) and UMC nurses and physicians; rank correlation with Hospital Anxiety Depression Scale (HADS) for anxiety (Anx) and depression are shown.

| Please mark on the line below to **show your worries** about the COVID-19 epidemic (0 to 100). That... | TH nurses |  |  |  |  | TH physicians |  |  |  |  | UMC nurses |  |  |  |  | UMC physicians |  |  |  |  |
| --- | --- | --- | --- | --- | --- | --- | --- | --- | --- | --- | --- | --- | --- | --- | --- | --- | --- | --- | --- | --- |
|  | Mean | SD | Median | Rank correlation (p-value) |  | Mean | SD | Median | Rank correlation (p-value) |  | Mean | SD | Median | Rank correlation (p-value) |  | Mean | SD | Median | Rank correlation (p-value) |  |
|  |  |  |  | Anx | Dep |  |  |  | Anx | Dep |  |  |  | Anx | Dep |  |  |  | Anx | Dep |
| I shall be infected | 58 | 29 | 54 | 0.33 | 0.26 | 59 | 26 | 61 | 0.02 | 0.003 | 63 | 30 | 71 | 0.45 | 0.26 | 57 | 30 | 57 | 0.49 | 0.37 |
|  |  |  |  | <.0001 | <.0001 |  |  |  | 0.83 | 0.97 |  |  |  | 0.0002 | 0.05 |  |  |  | 0.0003 | 0.01 |
| I shall infect my family | 72 | 29 | 81 | 0.30 | 0.20 | 73 | 27 | 81 | 0.08 | 0.09 | 70 | 30 | 79 | 0.49 | 0.32 | 72 | 30 | 80 | 0.48 | 0.32 |
|  |  |  |  | <.0001 | <.0001 |  |  |  | 0.38 | 0.31 |  |  |  | <.0001 | 0.01 |  |  |  | 0.001 | 0.02 |
| I shall infect my patients | 53 | 32 | 52 | 0.21 | 0.11 | 55 | 30 | 53 | 0.05 | 0.03 | 46 | 34 | 50 | 0.17 | 0.20 | 50 | 32 | 51 | 0.35 | 0.34 |
|  |  |  |  | <.0001 | 0.01 |  |  |  | 0.56 | 0.71 |  |  |  | 0.19 | 0.13 |  |  |  | 0.01 | 0.02 |
| I shall infect my co-workers/colleagues | 54 | 31 | 52 | 0.26 | 0.17 | 56 | 27 | 55 | 0.09 | 0.05 | 57 | 32 | 55 | 0.18 | 0.26 | 53 | 32 | 54 | 0.40 | 0.34 |
|  |  |  |  | <.0001 | <.0001 |  |  |  | 0.31 | 0.57 |  |  |  | 0.17 | 0.04 |  |  |  | 0.004 | 0.01 |
| I shall not be able to cope with the work | 42 | 31 | 38 | 0.42 | 0.32 | 34 | 28 | 27 | 0.33 | 0.33 | 43 | 35 | 44 | 0.39 | 0.32 | 34 | 29 | 28 | 0.30 | 0.21 |
|  |  |  |  | <.0001 | <.0001 |  |  |  | <.0001 | <.0001 |  |  |  | 0.002 | 0.01 |  |  |  | 0.04 | 0.14 |
| I shall have to let people die | 41 | 33 | 38 | 0.26 | 0.16 | 31 | 29 | 21 | 0.29 | 0.18 | 42 | 36 | 36 | 0.23 | 0.18 | 29 | 30 | 23 | 0.27 | 0.23 |
|  |  |  |  | <.0001 | <.0001 |  |  |  | 0.001 | 0.04 |  |  |  | 0.08 | 0.2 |  |  |  | 0.06 | 0.11 |
| my experience is inadequate | 34 | 30 | 25 | 0.33 | 0.18 | 31 | 27 | 21 | 0.26 | 0.17 | 34 | 31 | 24 | 0.16 | 0.25 | 24 | 22 | 20 | 0.34 | 0.30 |
|  |  |  |  | <.0001 | <.0001 |  |  |  | 0.003 | 0.05 |  |  |  | 0.2 | 0.05 |  |  |  | 0.02 | 0.03 |
| I shall fail myself and my family | 40 | 34 | 33 | 0.48 | 0.31 | 29 | 33 | 15 | 0.34 | 0.30 | 41 | 36 | 33 | 0.53 | 0.42 | 27 | 25 | 21 | 0.58 | 0.61 |
|  |  |  |  | <.0001 | <.0001 |  |  |  | <.0001 | 0.0004 |  |  |  | <.0001 | 0.001 |  |  |  | <.0001 | <.0001 |

### Multivariable models of HCWs at Tower Health

Adjusted effect estimates of covariates examined above for TH cohort are summarized in **Table VI**; effect estimates for perceptions is shown per 25 units (about one SD). Similar analyses for UMC did not produce stable models and therefore are not reported, although their findings largely agree with patterns seen at TH. After controlling for all other evaluated circumstances, the higher pneumonia symptom score (CAP-Sym) over a two-day period since diagnosis of the first COVID-19 case in the state was the most consistent predictor of higher risk of anxiety and depression across the two professions. Plots of observed and predicted HADS scores in relation to CAP-Sym suggest good model fit (**Appendix A**). This factor was correlated with belief in having been infected, which, being seen by us as an intermediate on the pathway towards anxiety and depression, was not forced into regression models; belief in having been infected is instead considered as pathway analysis below. For TH nurses, rank correlation of belief that they were infected with CAP-Sym was 0.4, p<0.0001, and for physicians -- 0.2, p=0.01.

**Table VI:**
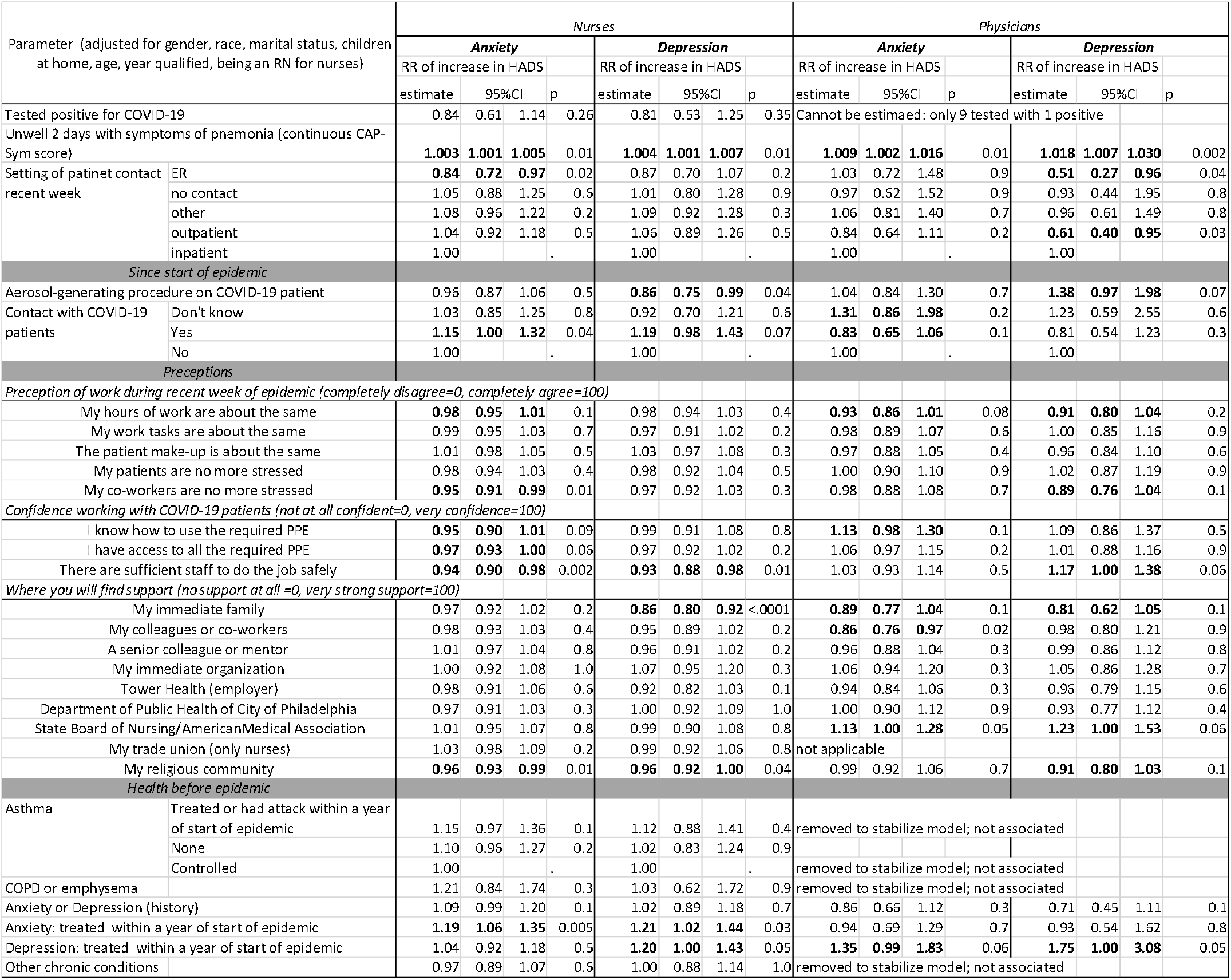
Adjusted associations with Hospital Anxiety Depression Scale (HADS) among Tower Health nurses (623) and physicians (153): negative binomial regression analysis with all listed variables included in the same model.

In adjusted analyses, nurses and physicians who recently encountered patients in emergency departments (ER) showed evidence of reduced risk of symptoms of anxiety and depression, respectively, relative to those who treated patients in the inpatient setting. Physicians who encountered patients in the outpatient settings were likewise less likely to show symptoms of depression relative to those who worked in inpatient settings. There was no evidence of other associations with setting of recent patient contact.

Having knowledge of any contact with COVID-19 patients was associated with, on average, 20% higher anxiety and depression scores in nurses relative to those who reported no such contact; no such associations were evident among physicians, except for a suggestion of reduced anxiety among physicians who thought than they had had contact with COVID-19 patients relative to those who did not (RR 0.83, 95%CI 0.65, 1.06). There was also some evidence that not knowing whether physicians encountered COVID-19 patients was a cause for anxiety (RR 1.31, 95%CI 0.86, 1.98). After allowing for knowledge of contact with COVID-19 patients, the reports of having performed aerosol-generating procedure on COVID-19 patients was not associated with anxiety but appeared to be related to reduced HADS scores for depression among nurses (RR 0.86, 95%CI 0.75, 0.99), with the opposite effect among physicians (RR 1.38, 95%CI 0.97, 1.98).

Among perceptions of work during most recent week of the epidemic, reports of working hours remaining the same and co-workers being no more stressed stood out as being associated with lower HADS scores, with the strongest effect estimate for lower anxiety scores among nurses who reported that their co-workers were “no more stressed” (RR 0.95, 95%CI 0.91, 0.99). Believing that there was enough staff to do the job safely was associated with reduced anxiety and depression scores among nurses only; anxiety was also lower among nurses who reported that they know how to use PPE and have access to it (with no such effect on the depression score). There was a suggestion that physicians who were confident in how to use PPE were also more anxious (RR 1.13, 95%CI 0.98, 1.30) and the those who were confident in having sufficient staff to do the job safely tended to be more depressed (RR 1.17, 95%CI 1.00, 1.38). Nurses and physicians who reported that they will have strong support from their families showed fewer symptoms of anxiety and depression, having allowed for all other factors in the analysis. Physicians who reported that they will find strong support from the American Medical Association were more likely to show symptoms of anxiety and depression; there was no analogous effect among nurses with respect to the State Board. Nurses who reported that they will find support in their religious community were less anxious and depressed; there was a suggestion of similar effect among physicians, especially for depression.

Among nurses, having been treated for anxiety a year before the epidemic was independently associated with HADS scores, with additional positive association between history of treatment for depression within a year of start of the epidemic and depression score. Among physicians, only report of history of recent pre-epidemic treatment for depression was associated with both higher anxiety and depression scores. No other elements of recorded medical histories appeared to independently relate to HADS scores.

After accounting for our measure of resilience did not materially alter the results despite its independent inverse association with anxiety and depression (details not shown).

### Path analyses

Results of path analyses for anxiety are summarized in **Figures 1** and **2**; excluding persons who tested positive for COVID-19 did not affect estimated associations. We did not examine all possible causal pathways, but merely estimated associations posited *a priori*, with only TH nurses, our largest sample, supplying evidence of existence of all hypothesized pathways (top of **Figure 1**). We estimated that a belief that a person was infected with COVID-19 (“Do you have reason to believe that you may have been infected with the COVID-19 virus?”: Yes/No) is directly related to higher HADS anxiety scores at both sites and professional groups. Likewise, the higher pneumonia (CAP-Sym) score was positively related to belief in having been infected. Data from TH revealed additional evidence of both the direct effect of CAP-Sym on anxiety and that mediated by belief in having been infected (**Figure 1**). Among TH nurses and physicians, and UMC physicians (but not nurses), there was a positive association between CAP-Sym and contact with COVID-19 patients: higher rate of history of pneumonia symptoms that lasted for two or more days was in those with contact with COVID-19 patients. Path analyses with depression score as outcome were different from those for anxiety in only two respects: there was no evidence that belief in having been infected was associated with HADS depression scores among UMC nurses and TH physicians, and there was evidence of direct effect of CAP-Sym on depression scores among UMC nurses. The results of these analyses are given in supplementals **Figures S1-S4 (Appendix B)**. Path analysis on data pooled across sites and professions yielded evidence of all hypothesized pathways (not shown).

**Figure 1:**
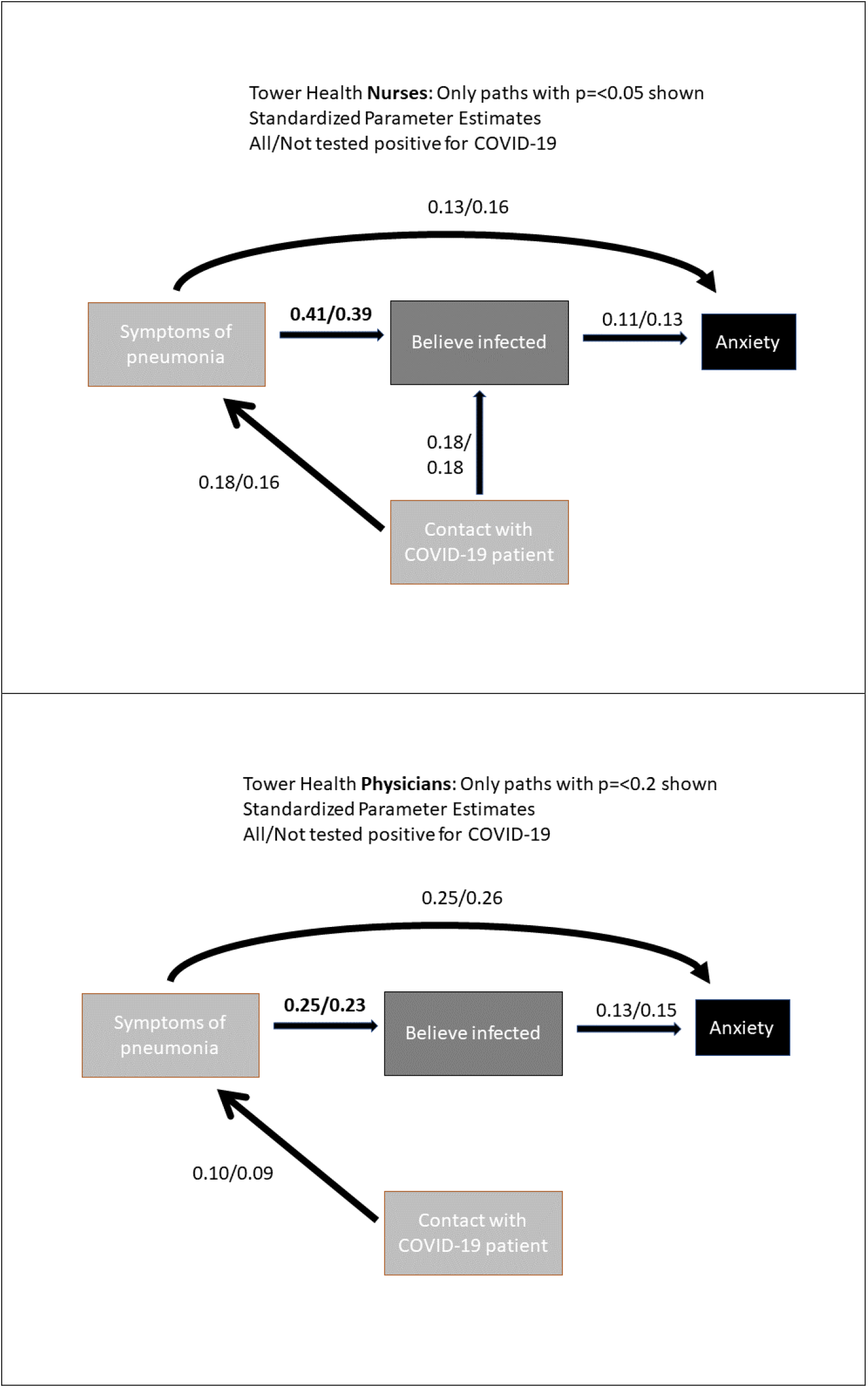
Pathways connecting Hospital Anxiety and Depression Scale **anxiety** score to symptoms of pneumonia (CAP-Sym) through belief of having been infected with virus that causes COVID-19, with consideration of contact with COVID-19 patients, among healthcare workers from Tower Health, PA (623 nurses (top) and 135 physicians (bottom)).

**Figure 2:**
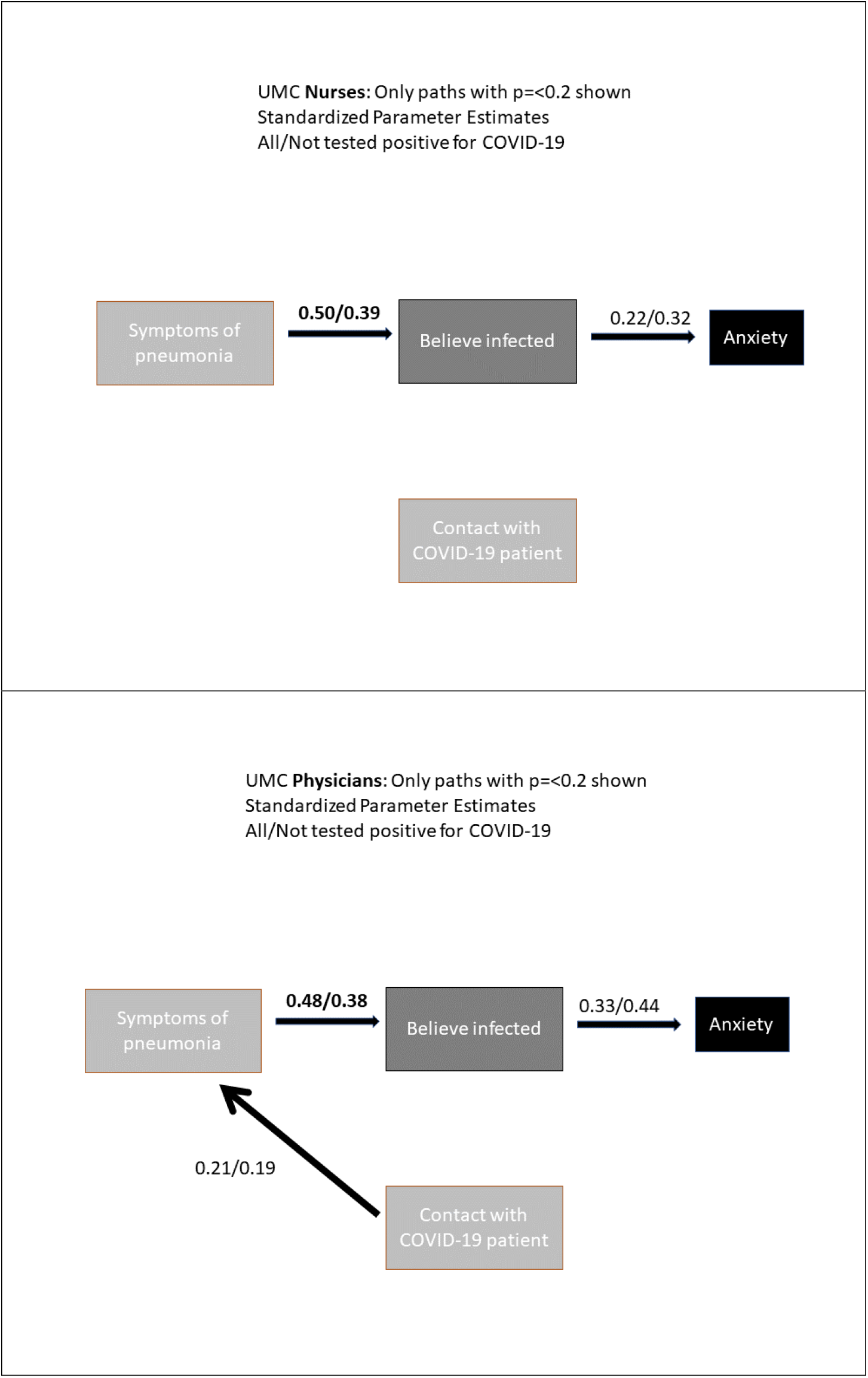
Pathways connecting Hospital Anxiety and Depression Scale anxiety score to symptoms of pneumonia (CAP-Sym) through belief of having been infected with virus that causes COVID-19, with consideration of contact with COVID-19 patients, among healthcare workers from the University Medical Center (UMC), Nevada, LV (61 nurses (top) and 50 physicians (bottom)).

## Discussion

We observed that about a third of nurses and physicians showed symptoms of anxiety or depression, which is similar, for the comparable time period, to findings by Czeisler et al.^17^ for US as the whole but lower than for the self-identified “essential workers” (42%). Differences in outcome assessment instruments make exact comparison problematic but HADS scores that we observed among HCWs are clearly above normative values established in the UK,^26^ with median normative scores for anxiety in 5-6 range and for depression about 3. We observed average scores greater than those reported by Cherry at al.^27^ for Canadian firefighters at the time they faced devastating Fort McMurray fires and were seen to develop elevated rates of post-traumatic stress disorder three years later. Specifically, we see evidence of greater than expected levels of anxiety and depression in nurses from both healthcare systems and physicians from TH but not UMC. This is likely an under-estimate of excess of prevalence of mood disorders among HCWs in the two healthcare systems because cross-sectional study design limited recruitment to active employees, excluding those who are too ill to work.

We evaluated resilience via the two-item Connor Davidson Resilience Scale but adjustment for it did not alter the results, suggesting that confounding by variation in “‘bounce-back’ and adaptability” in our samples is unlikely. The mean resilience scores were typical of US general population among physicians (7 out to maximum of 8), but disturbingly, in the range of family medicine and psychiatric outpatients among nurses (around 6).^21^ This is concordant with higher levels of symptoms of anxiety and depression among nurses, reinforcing the suggestion that mental health of nurses is more severely affected by the COVID-19 pandemic than that of physicians in the studied settings.

The most expressed worries were that of infection transmitted to the HCW and their family and oneself, with far fewer worries about performance of professional duties. Apprehension of failure of one’s own expectations of oneself and that of their family was the strongest correlate of anxiety and depression.

Although it is tempting to speculate that addressing these specific worries through mental health support services may have alleviated the burden of symptoms of anxiety and depression overall, it is not clear this would have alleviated either anxiety or depression.

Belief in having been infected (whether tested positive for the virus or not) emerged as a prominent cause of anxiety and depression, related more to history of symptoms known to HCWs to be consistent with COVID-19 at the time when testing may have been both limited and unreliable (not trusted), than to contact with infected patients. Among work-related factors that we identified as protective against anxiety and depression were having confidence in competent use and access to PPE, maintaining usual working hours and being surrounded by colleagues who were both sufficient in numbers and not stressed. Having support of immediate family and religious communities lessened anxiety and depression after accounting for other factors but any support was beneficial, although it was mostly believed that it will come from personal connections rather than professional bodies. There was some evidence that HCWs in emergency departments were less anxious and depressed and no clear evidence that involvement in aerosol-generating procedures on the infected patients was important per se.

Strengths of our work includes the use of HADS scores, which are more precise than commonly employed alternatives in large-scale epidemiologic studies,^28^ such as Patient Health Questionnaire (PHQ-4) used by Czeisler et al.^17^ However, it would have been desirable to employ measure of mood disorders that is directly comparable to the literature emerging from China and Canada. All previous work employed *ad hoc* questions of unknown psychometric properties to assess symptoms of COVID-19, while we used a validated questionnaire that captured symptoms by noting that they are consistent with community-acquired pneumonia. Thus, our analysis is less prone from bias due to errors in key outcomes and exposures.

Perception and concerns questions were developed specifically for our study and we did not have a chance, due to the punishing timetable imposed by the pandemic, to assess their reliability and validity. However, we are reassured by the fact that they yielded expected associations but acknowledge that bias from residual differential measurement error is possible. Differential measurement error may have arisen if, plausibly, persons more distressed by experience during epidemic were more likely to participate and made a greater effort in accurately responding to perceptions and concerns questions. Such selection mechanism may bias both internal and external validity of our findings and we are not able to address them quantitatively due to lack of information on even the demographics of non-participants. These concerns are aggravated by participation rate of 5-10%. However, our sample size is sufficient to yield robust inference (with adjustment for multiple factors via regression modeling) for the larger of the samples at TH and is informative of the situation experienced by selected participants at UMC. External validity of our findings is undermined by not including representative range of HCWs, such as licensed practical nurses, physician assistants, etc. However, existence of some concordance among studied professions among HCWs (mostly registered nurses and medical doctors) leaves us optimistic that some of the patterns we observe may be informative of the experience of all healthcare workers, the notion that is supported by our findings being largely in agreement with those from other jurisdictions.

There are likely factors related to working conditions (and their perceptions) and mood disorders that were not captured in our data, like insomnia and substance use, that could have confounded observed associations. However, we believe that we captured major confounders among our demographic and health-related variables, such that the risk of latent cofounding is reduced though regression adjustment for TH nurses and physicians. Measured confounders had little impact on direction and magnitude of the associations with pneumonia symptoms and associations with perceptions of PPE and working conditions, reassuring us in the robustness of these observations. We controlled for pre-existing mental health issues in isolating epidemic-related causes of anxiety and depression, further reducing the chance of bias in the results.

There was some heterogeneity in findings among two study sites, but they may be either due to chance or local peculiarities of healthcare systems’ and States response to the pandemic.

There were some differences in level of stress and anxiety and their correlates between nurses and physicians. This may be in part attributed to patient contact being typically is longer and more intimate for nurses. However, common themes also emerged, specifically related to pathway by which experience of pneumonia symptoms, contact with known COVID-19 patients, and belief in having been infected related to symptoms of anxiety and depression. There was no evidence that differences in anxiety and depression seen between nurses and physicians are explained by gender alone: results of regression analysis of pooled data adjusting for gender, profession and site are not shown, but revealed excess risk among nurses relative to physicians after accounting for gender.

We conclude that the levels and correlates of anxiety and depression among physicians and nurses in two US healthcare systems reveal that their experiences are like those of their colleagues around the world. It is not our place to speculate about specific mitigation measures that healthcare systems may wish to pursue to alleviate the burden of anxiety and depression among healthcare workers. Instead, we trust that our findings will help develop such measures and underscore the need to help nurses and physicians bear the psychological burden of combating COVID-19 pandemic and similar events in the future.

## Data Availability

Data is confidential as per IRB approvals.

## Acknowledgements

The authors are deeply indebted to all the participants who responded to survey while learning to live and work under disruptions precipitated by the pandemic. Dr Nicola M Cherry of the University of Alberta generously shared ideas and materials on related research. Staff of the two healthcare systems provided invaluable support in deploying the surveys.

## Notes

### Competing Interest Statement

The authors have declared no competing interest.

### Funding Statement

The authors report that there was no funding source for the work that resulted in the article or the preparation of the article.

### Author Declarations

Ethics approvals were obtained from Drexel University and the University of Nevada, Las Vegas for TH and UMC sites, respectively.

