## Appendix A for "A cross-sectional survey of the workplace factors contributing to symptoms of anxiety and depression among nurses and physicians during the first wave of COVID-19 pandemic in two US healthcare systems"

Appendix A: Predicted and observed HADS scores for anxiety and depression among Tower Health (TH) nurses and physicians in relation to pneumonia symptom (CAP-Sym) scores; dashed line is at 11, a cutoff for the “case” definition; regressions are least square linear regression on observed and predicted values

Nurses:

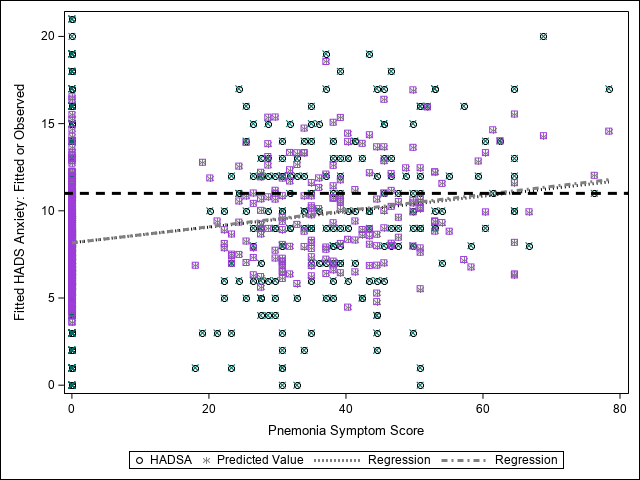

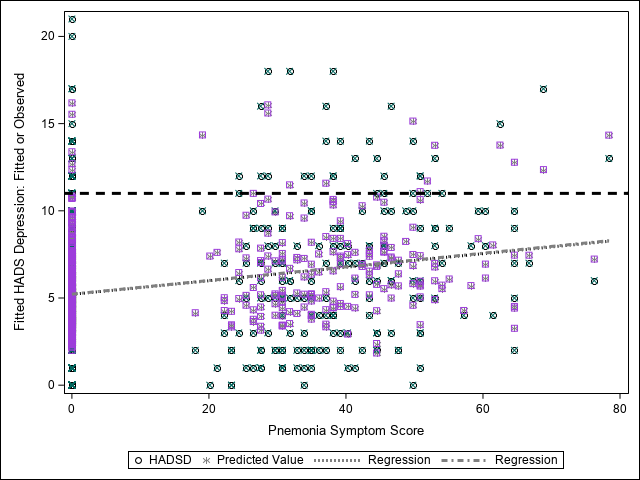

Physicians:

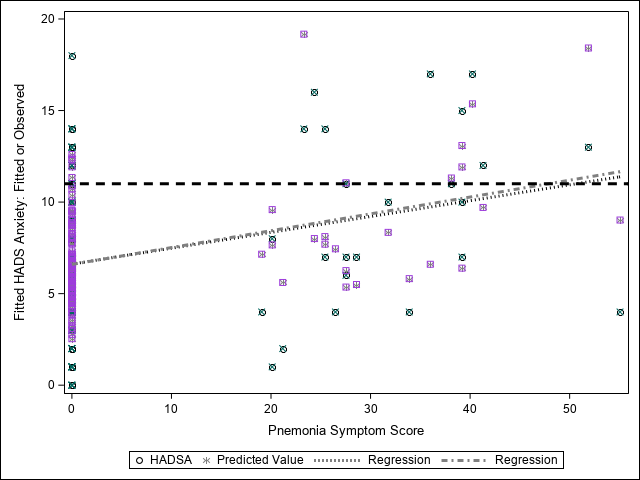

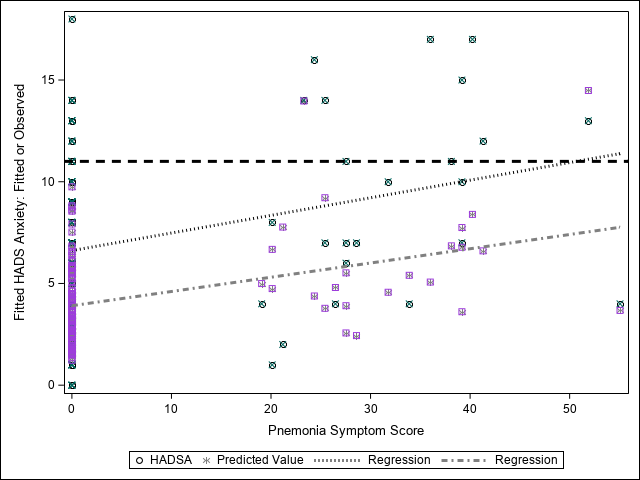
