## Appendix B for "A cross-sectional survey of the workplace factors contributing to symptoms of anxiety and depression among nurses and physicians during the first wave of COVID-19 pandemic in two US healthcare systems"

### Appendix B: Path analysis of HADS depression scores

**Figure S1:** Tower Health **Nurses**: Only paths with p=<0.05 shown
Standardized Parameter Estimates
All/Not tested positive for COVID-19

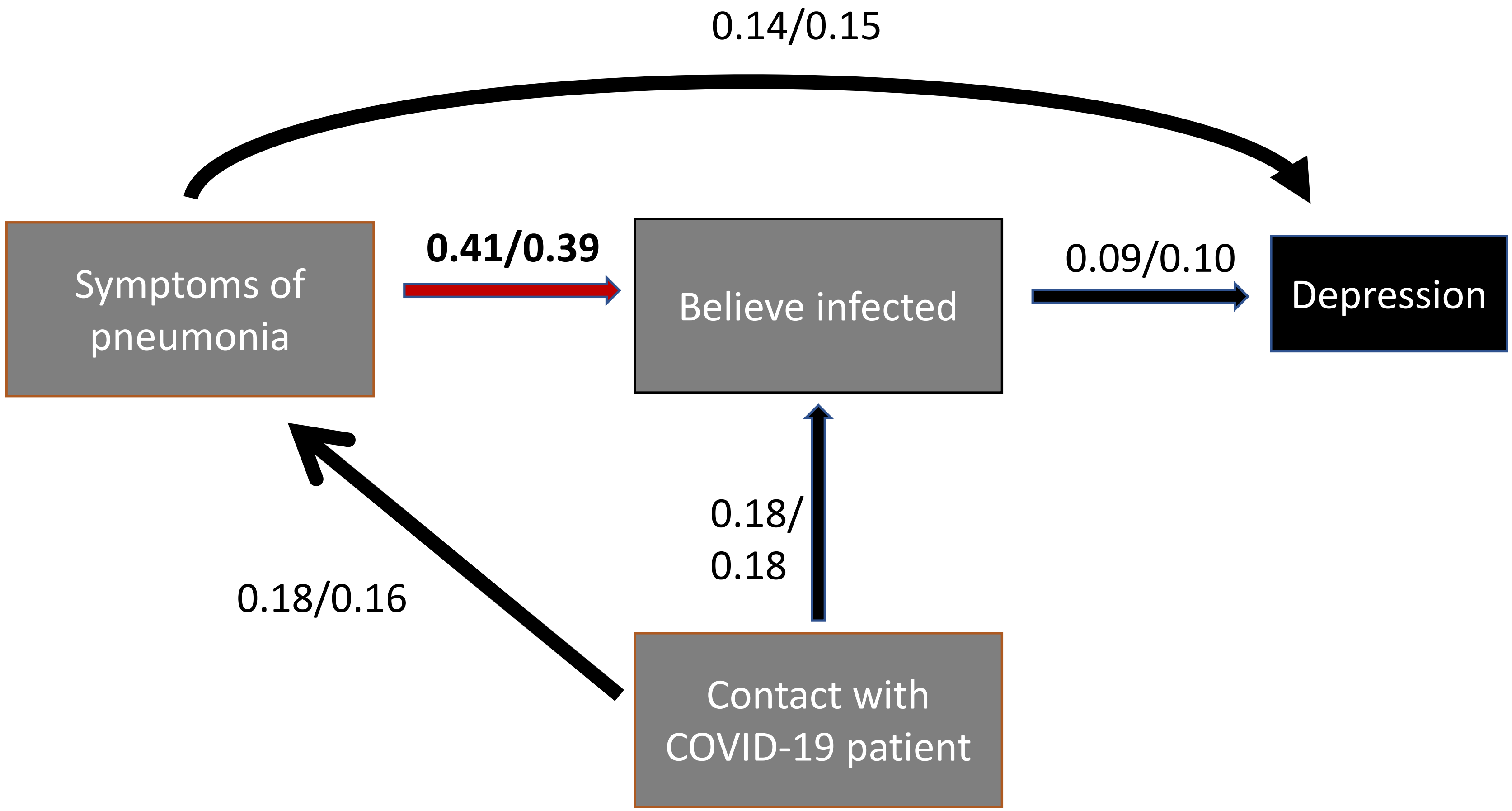

**Figure S2:** UMC **Nurses**: Only paths with p=<0.2 shown
Standardized Parameter Estimates
All/Not tested positive for COVID-19

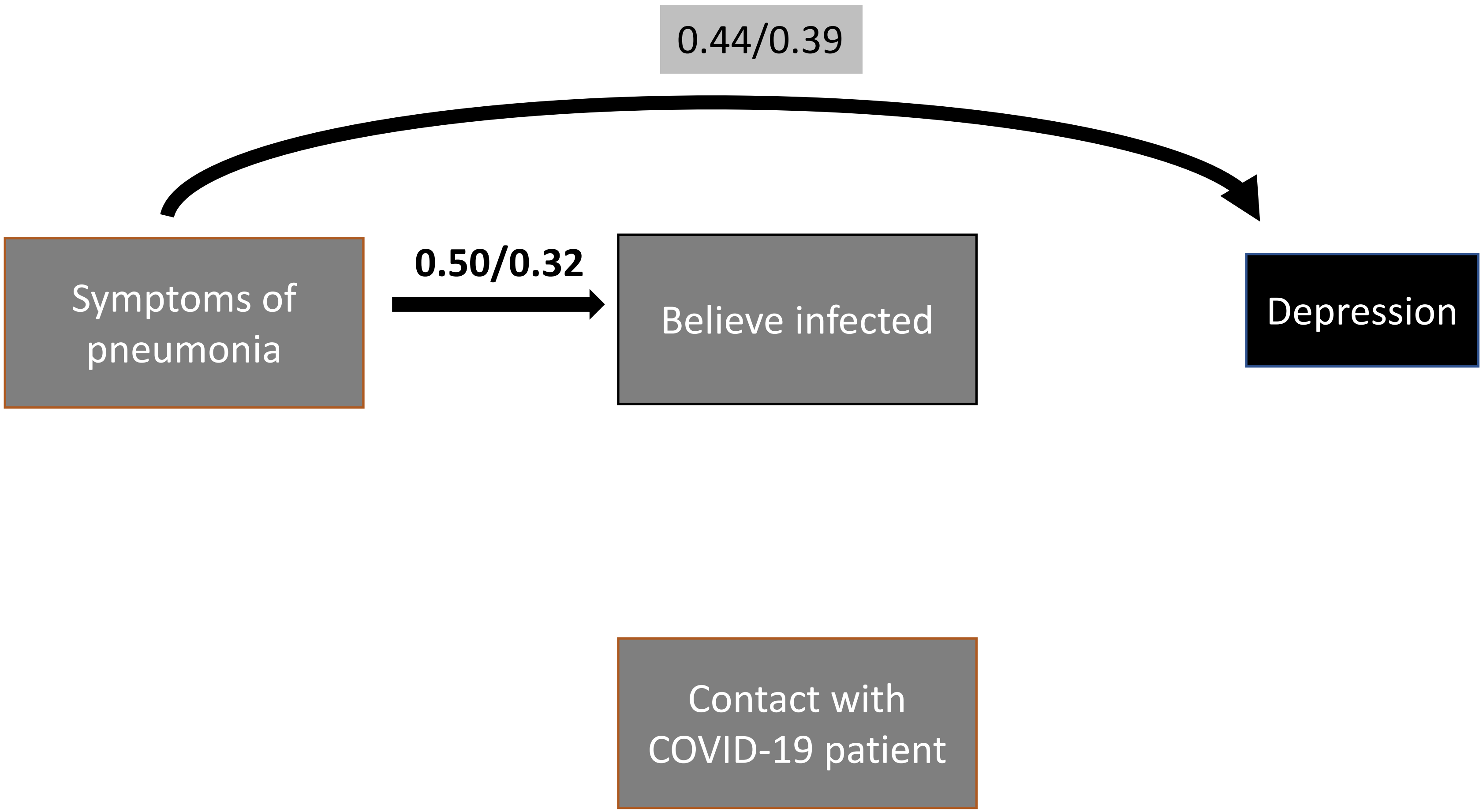

Figure S3: Tower Health **Physicians**: Only paths with p=<0.2 shown
Standardized Parameter Estimates
All/Not tested positive for COVID-19

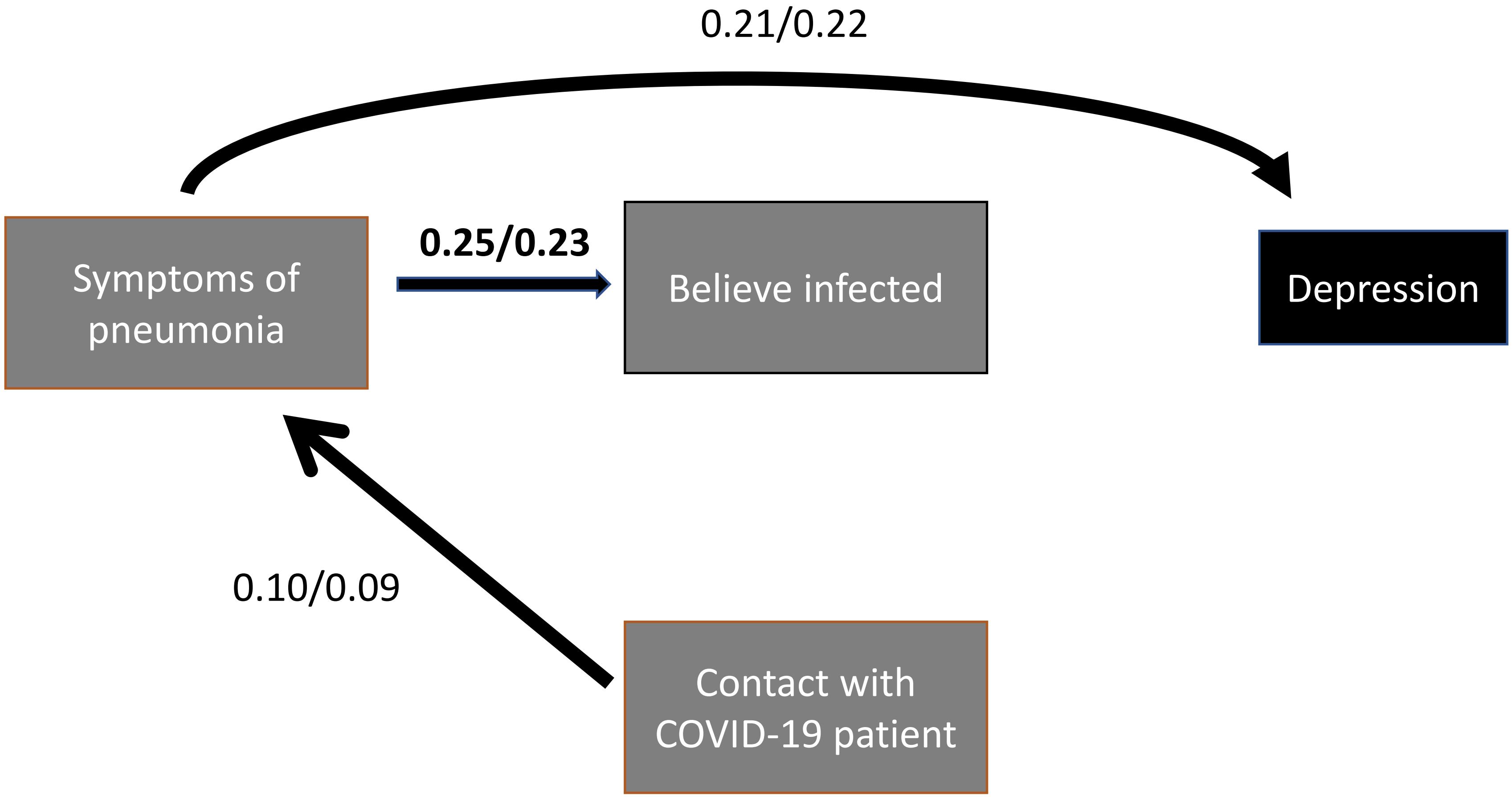

**Figure S4**: UMC **Physicians**: Only paths with p=<0.2 shown
Standardized Parameter Estimates
All/Not tested positive for COVID-19

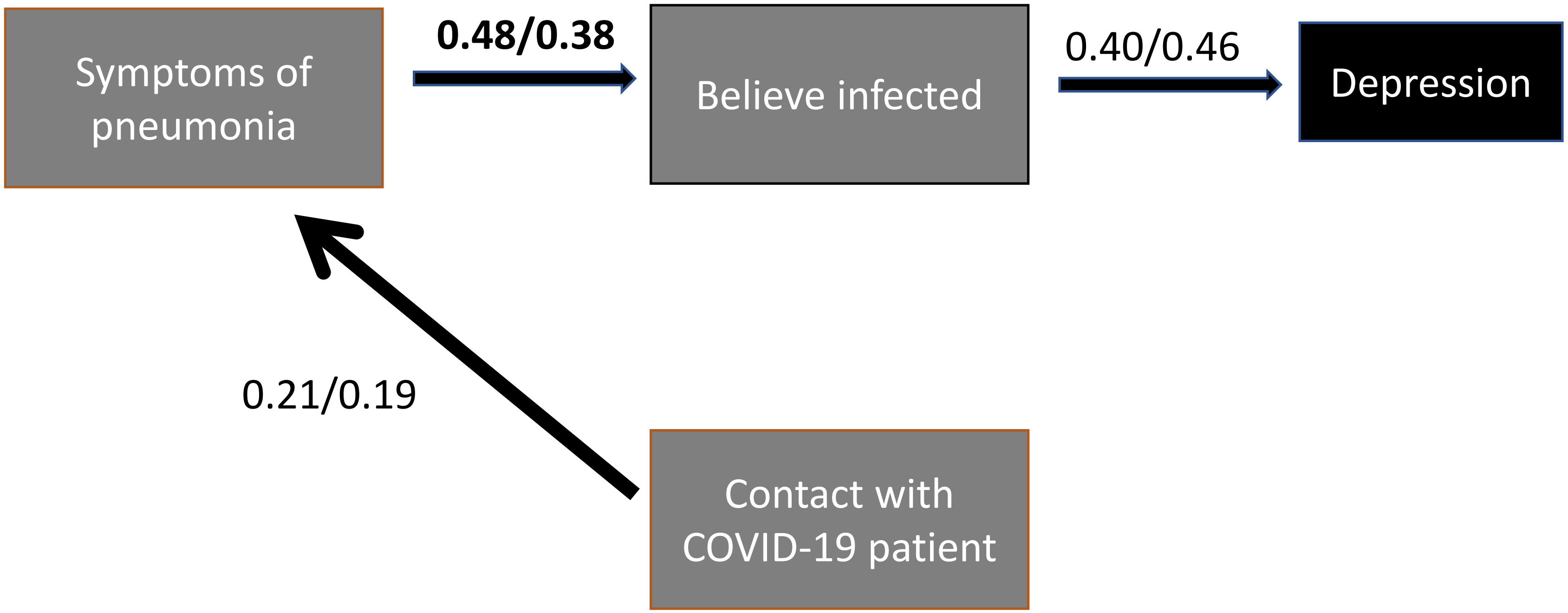
